# Implementation of school-feeding programme and its effect on nutritional adequacy among primary school children in Kisarawe district, Tanzania

**DOI:** 10.1101/2025.09.24.25335558

**Authors:** Julieth Sebba, Felistar Mwakasungura, Rebecca Mkumbwa, Bruno Sunguya

**Affiliations:** Department of Community Health, School of Public Health and Social Sciences, Muhimbili University of Health and Allied Sciences, 9, United Nations Road, Upanga West 11103, Dar es salaam, Tanzania; Department of Community Health, School of Public Health, KCMC University, Moshi, Tanzania; Department of Developmental Studies, School of Public Health and Social Sciences, Muhimbili University of Health and Allied Sciences, 9, United Nations Road, Upanga West 11103, Dar es salaam, Tanzania

**Keywords:** School Feeding Program, Dietary Diversity, Meal Frequency, Nutritional Adequacy

## Abstract

The school-age period is crucial nutritional-wise owing to its health and developmental impact. In Tanzania, 11% of school-aged children are thin, while 34% are anemic, representing a significant burden of undernutrition with a potential to have a lasting impact on educational and early onset of non-communicable diseases. This study was designed to evaluate its implementation and effects on nutritional adequacy among primary-school children in Kisarawe District, Tanzania.

A school-based cross-sectional study was conducted among 300 students aged 10-14 years from 10 primary schools from March to May 2025. Nutritional adequacy was assessed using two measures: Dietary diversity and Meal frequency. Descriptive statistics was used to summarize the status of implementation of the school feeding program, while regression analysis was used to identify the effect of the intervention.

All ten schools implemented some components of the school feeding program with a significant limitation. Only 65% students were covered in the intervention. Forty-nine percent of students met the minimum dietary diversity score. Participation in a school feeding program was associated with a 45% higher likelihood of attaining minimum meal diversity (APR = 1.45; 95% CI: 1.00-2.12; p = 0.051), although this did not reach statistical significance threshold. Only 15.3% students reported consuming the minimum of three meals per day.

School feeding programs were present in Kisarawe district but faced major infrastructural and logistical challenges, with none of the schools fully adhering to national guidelines. Strengthening infrastructure, ensuring reliable food supply, and expanding universal coverage are essential to maximize program impact.

## Introduction

Proper nutrition during school years is crucial, as it significantly enhances children’s ability to grow, learn, and thrive (1). Adequate nutrition during these critical years can enable catch-up growth for children who have suffered from malnutrition in early childhood, with some effects on detrimental impact of earlier nutritional deficits (2). Nutritional adequacy can primarily be assessed through dietary diversity and meal frequency. Higher dietary diversity and meal frequency improve micronutrient intake and the prevention of chronic effects of undernutrition such as anemia and stunting (3–6).

Globally, severe undernutrition affects approximately 45 million children annually, while 149 million school-aged children live with thinness, highlighting a substantial nutritional burden worldwide (7,8). Sub-Saharan Africa faces particularly severe nutritional challenges driven by pervasive poverty, conflict, climate crises, and a high prevalence of infectious diseases, exacerbating the nutritional vulnerabilities of school-aged children in this region (9–11). Tanzania is similarly challenged, with 11% of school-aged children classified as thin and 34% suffering from anemia (12). These nutritional deficiencies compromise children’s health, educational outcomes, and future socio-economic potential.

The adverse effects of severe undernutrition during school years extend beyond immediate physical impacts to long-term cognitive and academic consequences. Undernutrition impairs cognitive and motor development, negatively influencing children’s capacity to learn effectively, thus leading to reduced academic performance and future productivity (13). Furthermore, malnourished children exhibit increased susceptibility to diseases due to weakened immunity, further compromising their overall health (14).

To address these nutritional challenges, Tanzania launched the National Guideline on School Feeding and Nutrition Services in 2021, providing structured guidance for implementing school feeding programs to improve nutritional adequacy among school-aged children (15). Additionally, Tanzania’s National Nutrition Plan 2025/2026 highlights the country’s objective of expanding school feeding programs through collaborative partnerships among local communities, ministries, and development organizations (12,16).

Despite these initiatives, undernutrition continues to be a pressing issue in Tanzania, particularly in areas like Kisarawe District, where nutritional deficiencies remain prevalent. Previous research indicates that anemia affects 53.3% of school-aged children in Tanzania’s Pwani region, including Kisarawe District, primarily due to limited dietary diversity characterized by high starch consumption and inadequate fruit and dairy intake (12). Some children are also coming from households with food insecurity, making food programs at school the sole solution for their daily nutritional adequacy pathway (17,18).

Kisarawe is among several Tanzanian districts currently implementing school feeding programs; however, the effects of these interventions in improving children’s nutritional adequacy remains inadequately documented (19). This study therefore aimed to examine the effect of school feeding programs on dietary adequacy measured through dietary diversity and meal frequency among school-aged children in Kisarawe District, Tanzania.

## Materials and methods

### Study design and area

This study was a school-based cross-sectional, employing a quantitative approach, it was conducted from March to May 2025 in Kisarawe District, Tanzania. The district, located in the Pwani region, is predominantly rural and relies heavily on agriculture. Main food crops from this district include cassava, maize, rice, sorghum, and sweet potatoes. The district was purposefully selected since it’s one of the districts in the Pwani Region, experiencing a high prevalence of chronic undernutrition among school-aged children.

### Study population and sampling

The study targeted school-aged children between 10 and 14 years enrolled in primary schools in Kisarawe District. A multi-stage sampling procedure was employed. Initially, wards were stratified into urban and rural clusters. Two wards from each cluster were conveniently selected to represent geographic diversity, resulting in a total of four wards. Within these wards, ten primary schools were chosen conveniently, five from urban and five from rural wards. From each school, simple random sampling using random number generators was used to select 300 participants, ensuring representativeness and reducing selection bias. Participants were selected from standard five and six to avoid disturbing classes four and seven from exam preparations, and lower classes were excluded to minimize recall bias.

### Variables and measurements

The exposure variable was participation in the school feeding program. The outcome variable was nutritional adequacy measured through attaining a minimum dietary diversity and meal frequency. Dietary diversity was assessed based on consumption of at least four out of seven predefined food groups: grains, legumes and nuts, dairy products, vitamin A-rich fruits and vegetables, other vegetables, meat and fish, and eggs within the previous 24 hours. Meal frequency was measured by the number of meals consumed within the previous 24 hours, with regular meal frequency defined as consumption of three or more meals (20). Covariates and confounders included demographic characteristics; parental characteristics including education levels and parental occupations; and household characteristics including socio-economic status, household size, and households’ food security status.

### Data collection

Data collection was conducted using structured questionnaires administered through face-to-face interviews in Swahili, facilitating comprehension and accuracy of responses. Dietary diversity and meal frequency were evaluated using a 24-hour dietary recall method. Participants detailed foods consumed from waking until bedtime the previous day. Dietary diversity scores were then calculated based on reported consumption across the seven food groups, with higher scores indicating greater dietary diversity. Meal frequency data were also captured during these interviews.

### Data management and analysis

Data were analyzed using Stata Version 15 software. Implementation status was calculated using a frequency table and findings were summarized to show the percentage of schools having the required items for the implementation of the school feeding program. Minimum Dietary diversity was calculated using a frequency table to get scores. Multilevel modified Poisson regression was used to determine the association between students’ participation in the school feeding program and minimum dietary diversity, whereby multivariate models was employed to determine the estimates of the crude odds ratio, variables with p<0.2 were used to find the adjusted prevalence ratio (APR) at 95% Confidence level. For meal frequency was obtained from a frequency table then Multilevel modified Poisson regression was used to determine the association between students’ participation in the school feeding program and meal frequency.

### Ethical approval, informed consent and Patient Participation

Ethical clearance was obtained from the Muhimbili University of Health and Allied Sciences Research Ethics Committee, IRB number: MUHAS-REC-03-2025-2693. Additional permissions were secured from district educational authorities and participating schools. Written informed consent was obtained from head teachers, and verbal assent from participating students. Confidentiality, voluntary participation, and data privacy were rigorously maintained, adhering to ethical standards throughout the study.

Patients and the public were not involved in the design, conduct, reporting, or dissemination of this study. They were engaged solely as participants.

## Results

### Socio-demographic characteristics of participants

A total of 300 school-aged children participated in this study, with ages ranging from 10 to 14 years. Their mean age was 11.5 (±1.1) years. The majority of parents or guardians had low educational attainment, with 204 (68%) of fathers and 218 (72.7%) of mothers having either no formal education or primary school education only. Food insecurity was reported by 42 (14%) of households, indicating limited access to adequate food within the previous month “Table 1”.

**Table 1:**
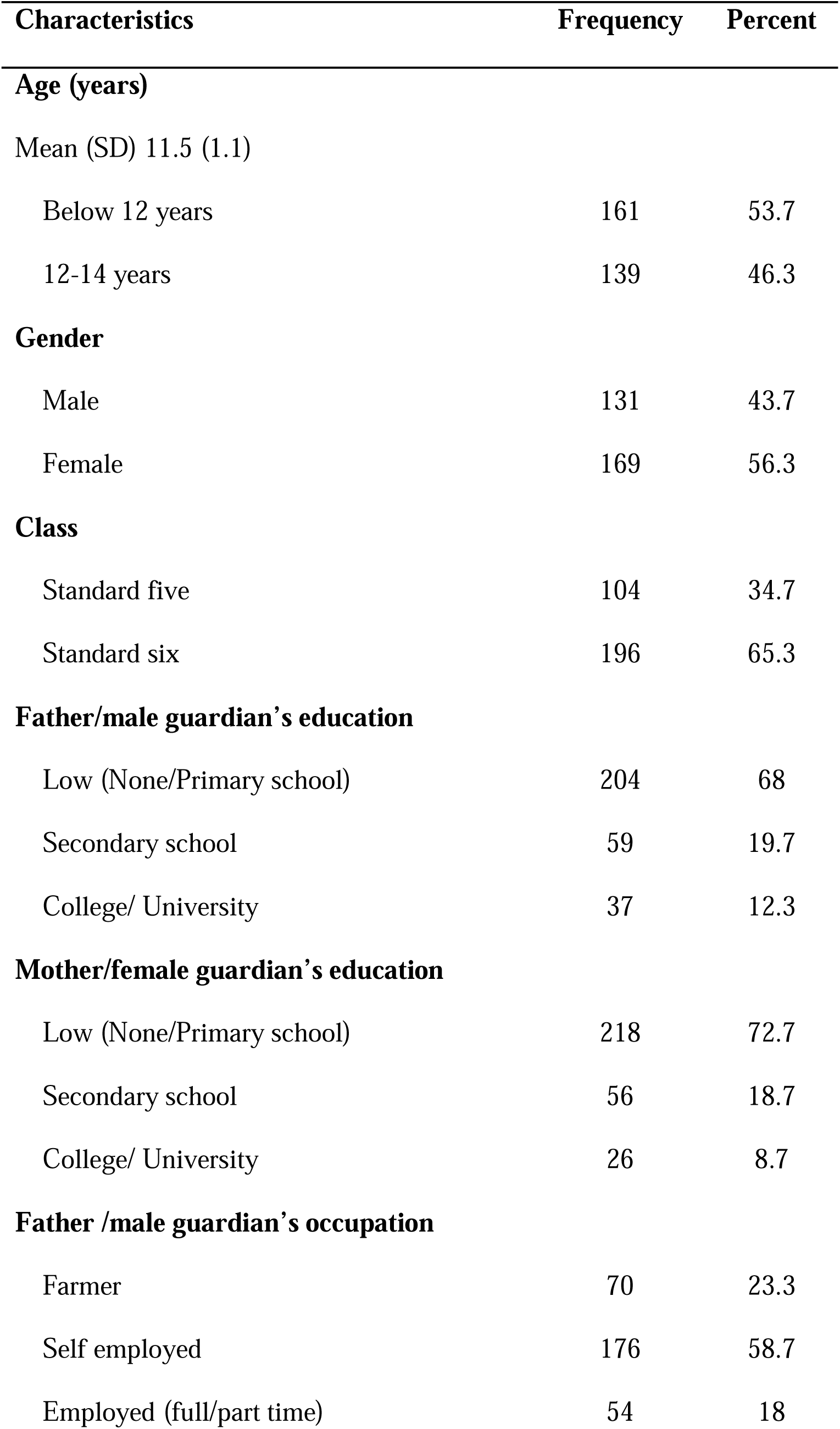
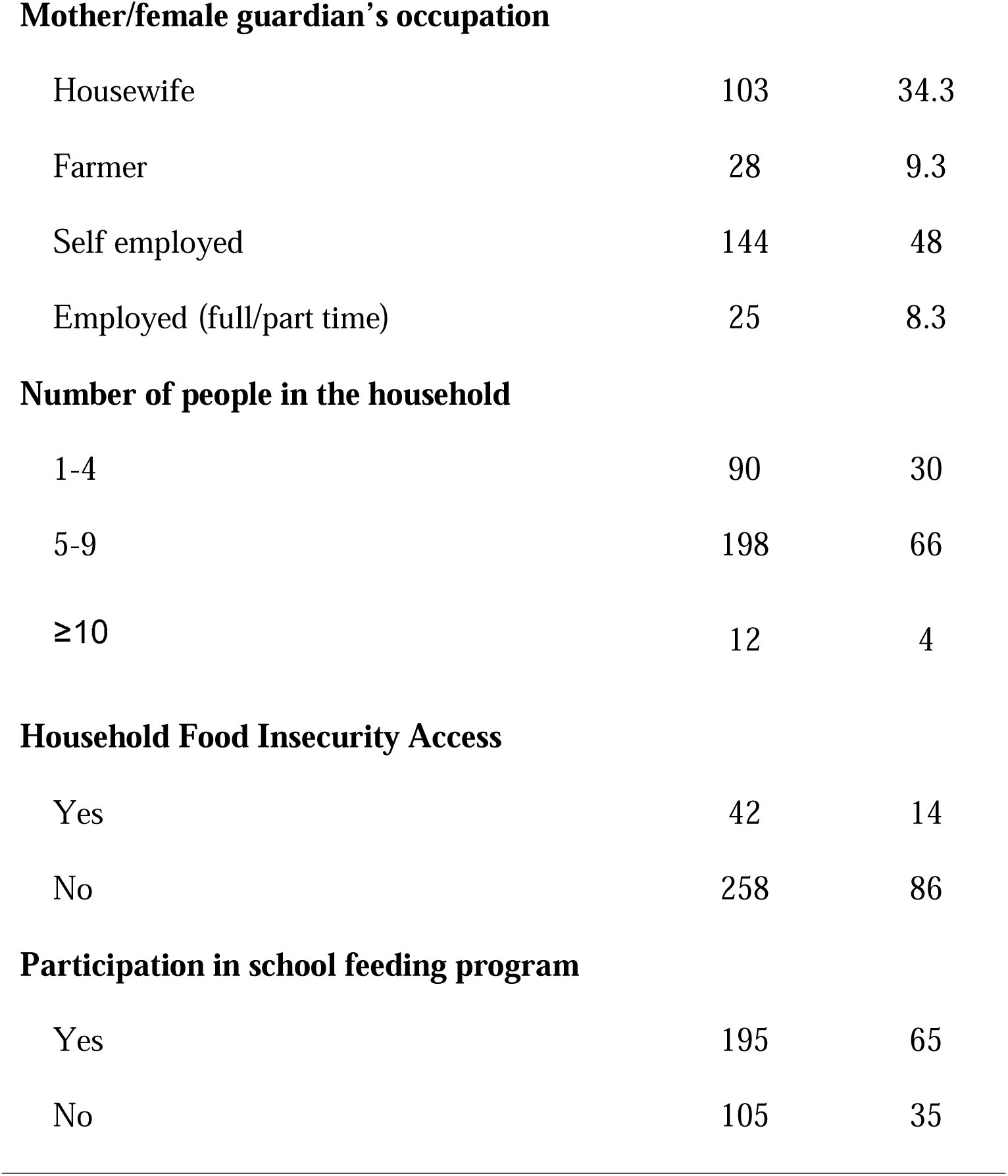
Socio-demographic characteristics of study participants (n=300).

| Characteristics | Frequency | Percent |
| --- | --- | --- |
| **Age (years)** |  |  |
| Mean (SD) 11.5 (1.1) |  |  |
| Below 12 years | 161 | 53.7 |
| 12-14 years | 139 | 46.3 |
| **Gender** |  |  |
| Male | 131 | 43.7 |
| Female | 169 | 56.3 |
| **Class** |  |  |
| Standard five | 104 | 34.7 |
| Standard six | 196 | 65.3 |
| **Father/male guardian's education** |  |  |
| Low (None/Primary school) | 204 | 68 |
| Secondary school | 59 | 19.7 |
| College/ University | 37 | 12.3 |
| **Mother/female guardian's education** |  |  |
| Low (None/Primary school) | 218 | 72.7 |
| Secondary school | 56 | 18.7 |
| College/ University | 26 | 8.7 |
| **Father /male guardian's occupation** |  |  |
| Farmer | 70 | 23.3 |
| Self employed | 176 | 58.7 |
| Employed (full/part time) | 54 | 18 |
| **Mother/female guardian's occupation** |  |  |
| Housewife | 103 | 34.3 |
| Farmer | 28 | 9.3 |
| Self employed | 144 | 48 |
| Employed (full/part time) | 25 | 8.3 |
| **Number of people in the household** |  |  |
| 1-4 | 90 | 30 |
| 5-9 | 198 | 66 |
| ≥10 | 12 | 4 |
| **Household Food Insecurity Access** |  |  |
| Yes | 42 | 14 |
| No | 258 | 86 |
| **Participation in school feeding program** |  |  |
| Yes | 195 | 65 |
| No | 105 | 35 |

### Implementation Status of School Feeding Programs

All ten schools surveyed had ongoing school feeding programs; however, none fully met the implementation criteria outlined in Tanzania’s National Guideline for School Feeding Programs. Notably, all schools lacked designated eating areas or cafeterias. Only 50% of schools had food storage facilities, whereas 70% possessed adequate cooking facilities. Additionally, 70% of schools consistently provided meals throughout the preceding academic year, but only 40% provided meals containing at least four different food groups as recommended. Despite the presence of school feeding programs, only 65% of enrolled students participated regularly due to economic barriers “Figure 1”.

**Figure 1:**
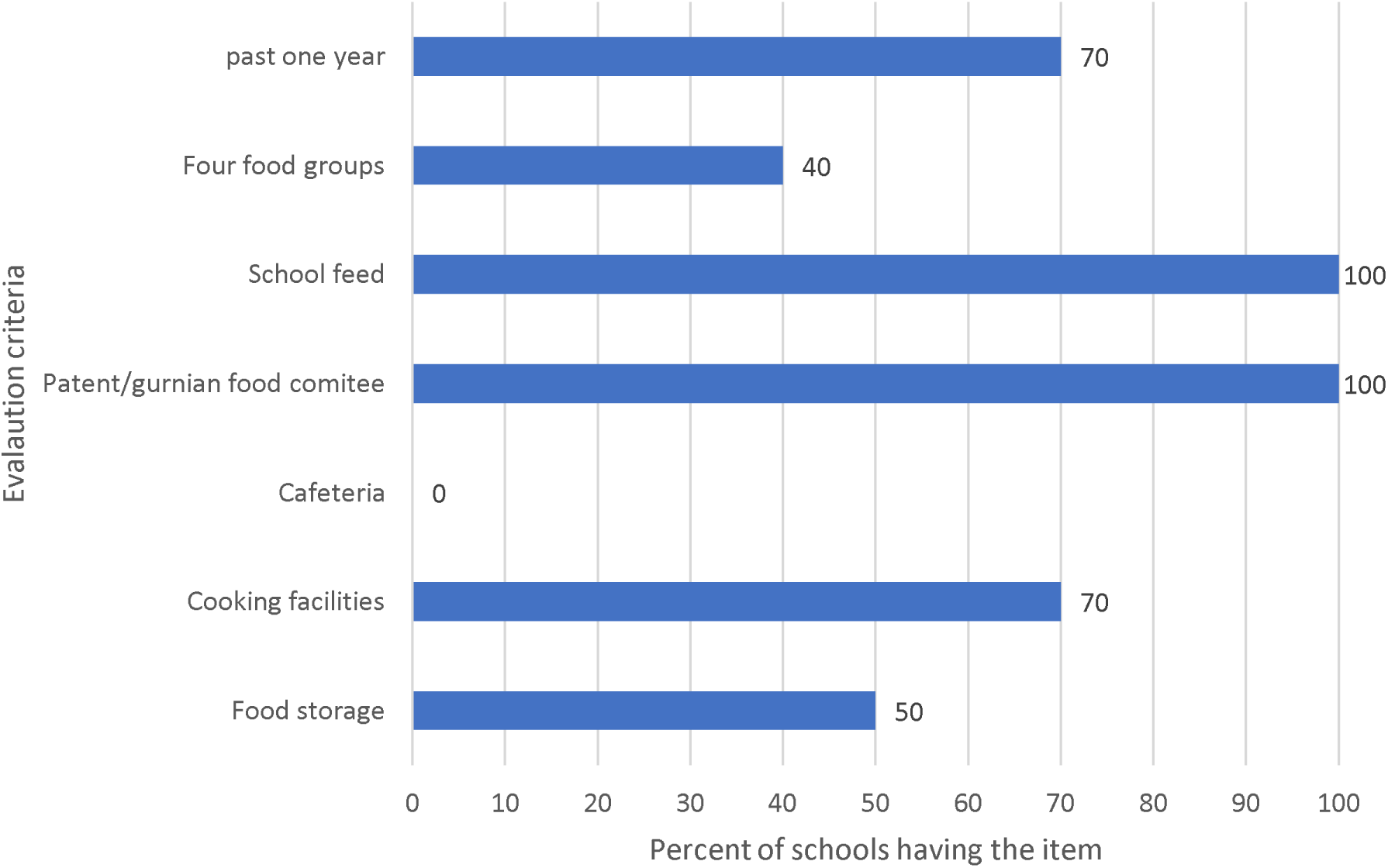
Availability of required items in schools to show implementation status.

### Association Between School Feeding Participation and Dietary Diversity

Nearly half of the enrolled students (49.3%) achieved minimum dietary diversity, defined as consuming at least four food groups within the previous 24 hours. Among those who achieved minimum dietary diversity, 74% reported participating in school feeding program.

Although participation in a school feeding program increase the likelihood of attaining MMD by 45%, the difference did not reach a statistically significance level (APR=1.45; 95% CI=1.00-2.12; p=0.051). Household food insecurity significantly reduced the likelihood of achieving minimum dietary diversity (APR=0.44; 95% CI=0.23-0.85; p=0.014).). Other socio-demographic variables, such as age, parental education and household size, showed no statistically significant association with dietary diversity after adjustment “Table 2”.

**Table 2:**
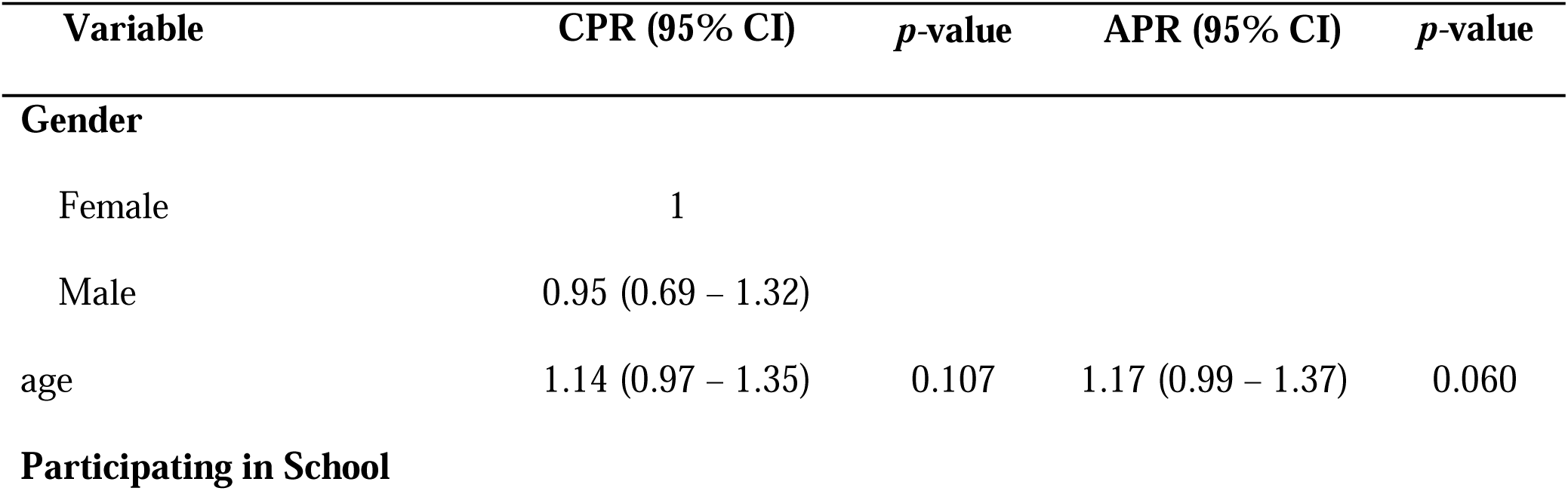
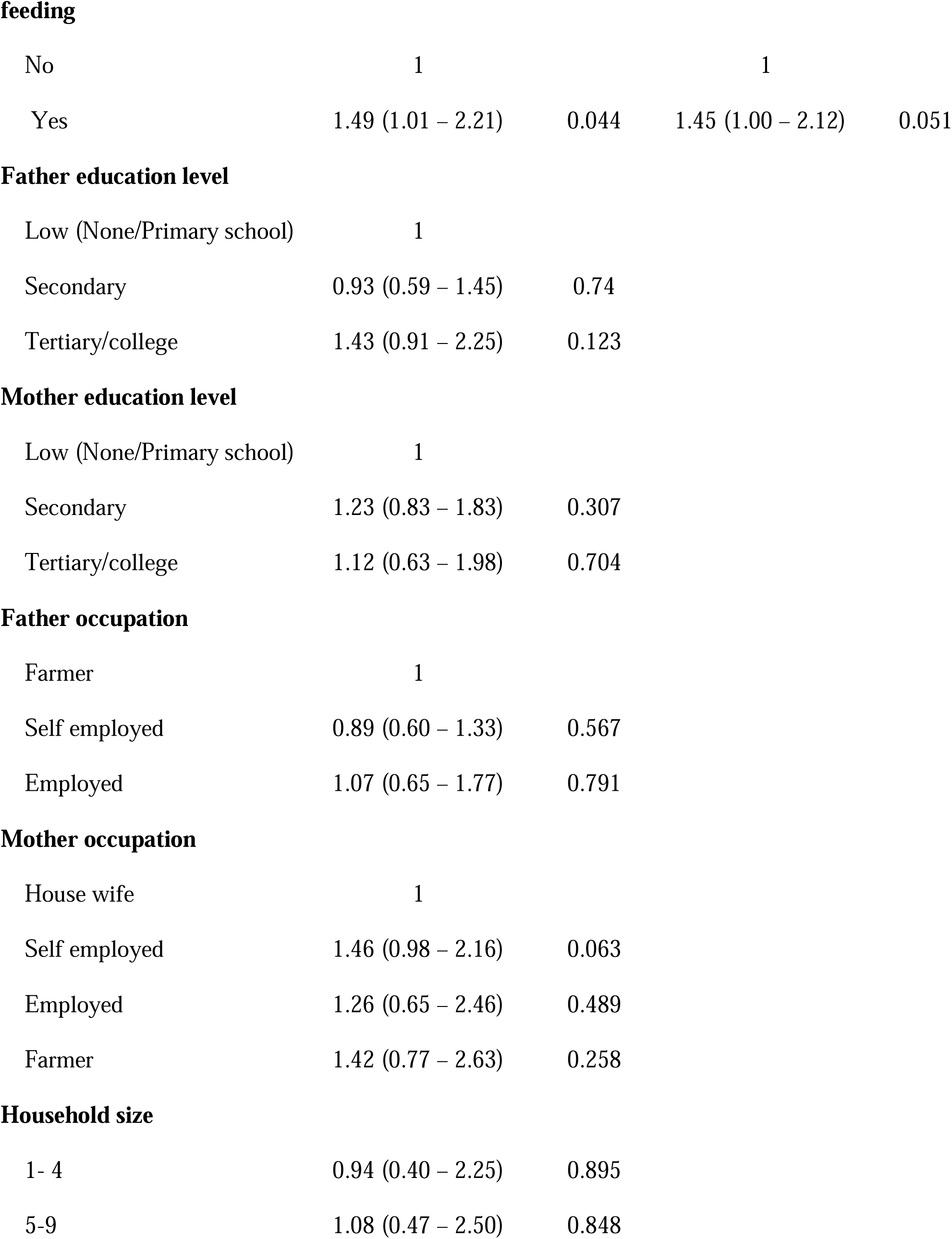
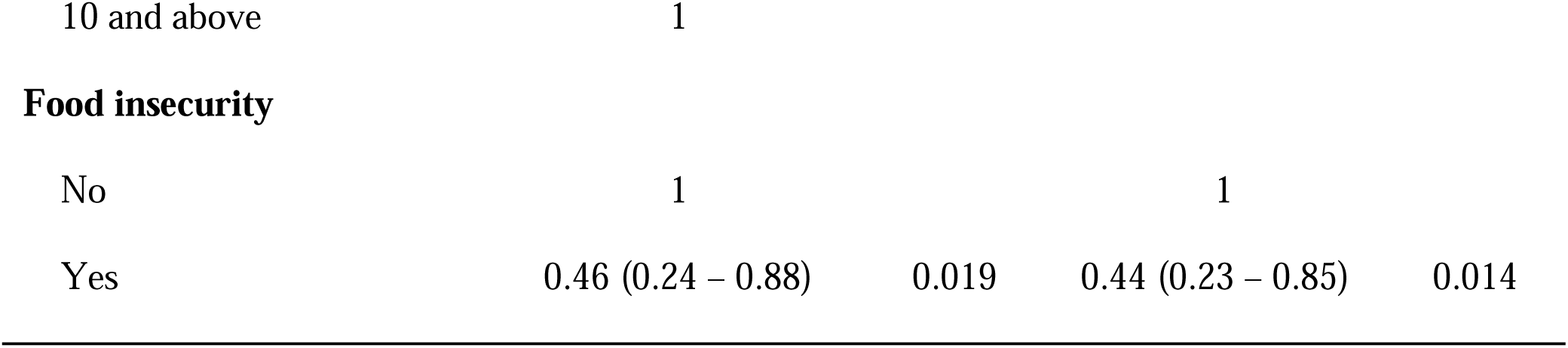
Association between participating in school feeding program and minimum dietary diversity.

| **Variable** | **CPR (95% CI)** | **p-value** | **APR (95% CI)** | **p-value** |
| --- | --- | --- | --- | --- |
| **Gender** |  |  |  |  |
| Female | 1 |  |  |  |
| Male | 0.95 (0.69 – 1.32) |  |  |  |
| age | 1.14 (0.97 – 1.35) | 0.107 | 1.17 (0.99 – 1.37) | 0.060 |
| **Participating in School** |  |  |  |  |
| **feeding** |  |  |  |  |
| No | 1 |  | 1 |  |
| Yes | 1.49 (1.01 – 2.21) | 0.044 | 1.45 (1.00 – 2.12) | 0.051 |

**Father education level**
|  |  |  |
| --- | --- | --- |
| Low (None/Primary school) | 1 |  |
| Secondary | 0.93 (0.59 – 1.45) | 0.74 |
| Tertiary/college | 1.43 (0.91 – 2.25) | 0.123 |

**Mother education level**
|  |  |  |
| --- | --- | --- |
| Low (None/Primary school) | 1 |  |
| Secondary | 1.23 (0.83 – 1.83) | 0.307 |
| Tertiary/college | 1.12 (0.63 – 1.98) | 0.704 |

**Father occupation**
|  |  |  |
| --- | --- | --- |
| Farmer | 1 |  |
| Self employed | 0.89 (0.60 – 1.33) | 0.567 |
| Employed | 1.07 (0.65 – 1.77) | 0.791 |

**Mother occupation**
|  |  |  |
| --- | --- | --- |
| House wife | 1 |  |
| Self employed | 1.46 (0.98 – 2.16) | 0.063 |
| Employed | 1.26 (0.65 – 2.46) | 0.489 |
| Farmer | 1.42 (0.77 – 2.63) | 0.258 |

**Household size**
|  |  |  |
| --- | --- | --- |
| 1- 4 | 0.94 (0.40 – 2.25) | 0.895 |
| 5-9 | 1.08 (0.47 – 2.50) | 0.848 |
| 10 and above | 1 |  |  |  |
| **Food insecurity** |  |  |  |  |
| No | 1 |  | 1 |  |
| Yes | 0.46 (0.24 – 0.88) | 0.019 | 0.44 (0.23 – 0.85) | 0.014 |

### Effect of School Feeding Participation on Meal Frequency

Overall, only 15.3% of enrolled students reported consuming the recommended minimum of three meals per day, inclusive of snacks. Among those who achieved minimum dietary diversity, 96% reported participating in school feeding program. Multivariable modified Poisson regression analyses further supported this association, revealing that students participating in the school feeding programs were nearly 12 times more likely to meet the recommended meal frequency than non-participants (APR=11.68; 95% CI=2.81-48.53; p=0.001). Gender also emerged as a significant factor, with males being 48% less likely to consume regular meals compared to females (APR=0.52; 95% CI=0.28-0.99; p=0.047). Other demographic variables, including parental occupation, education level, household size, and food insecurity, did not show significant associations with meal frequency in the adjusted model “Table 3”.

**Table 3:**
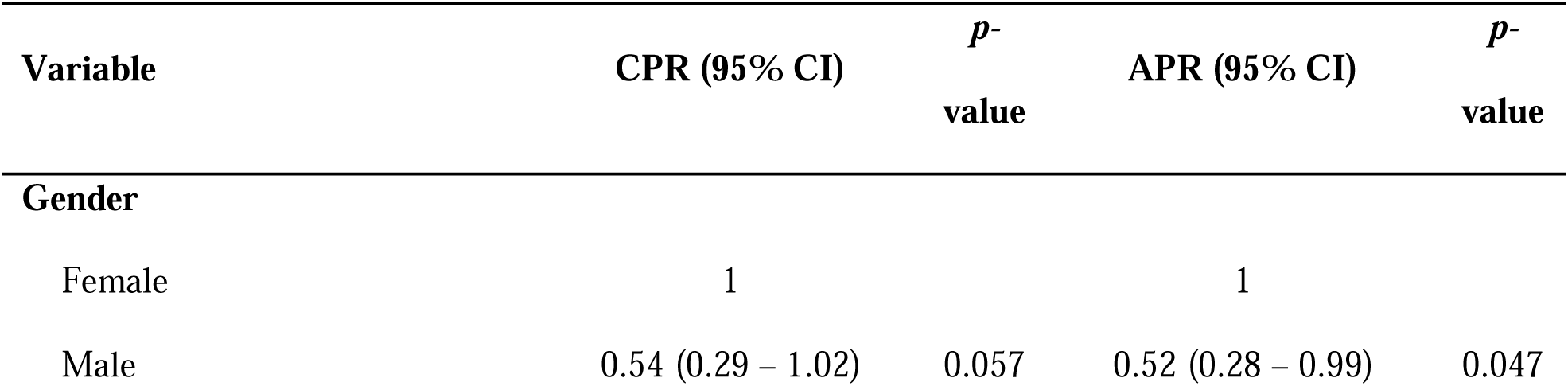
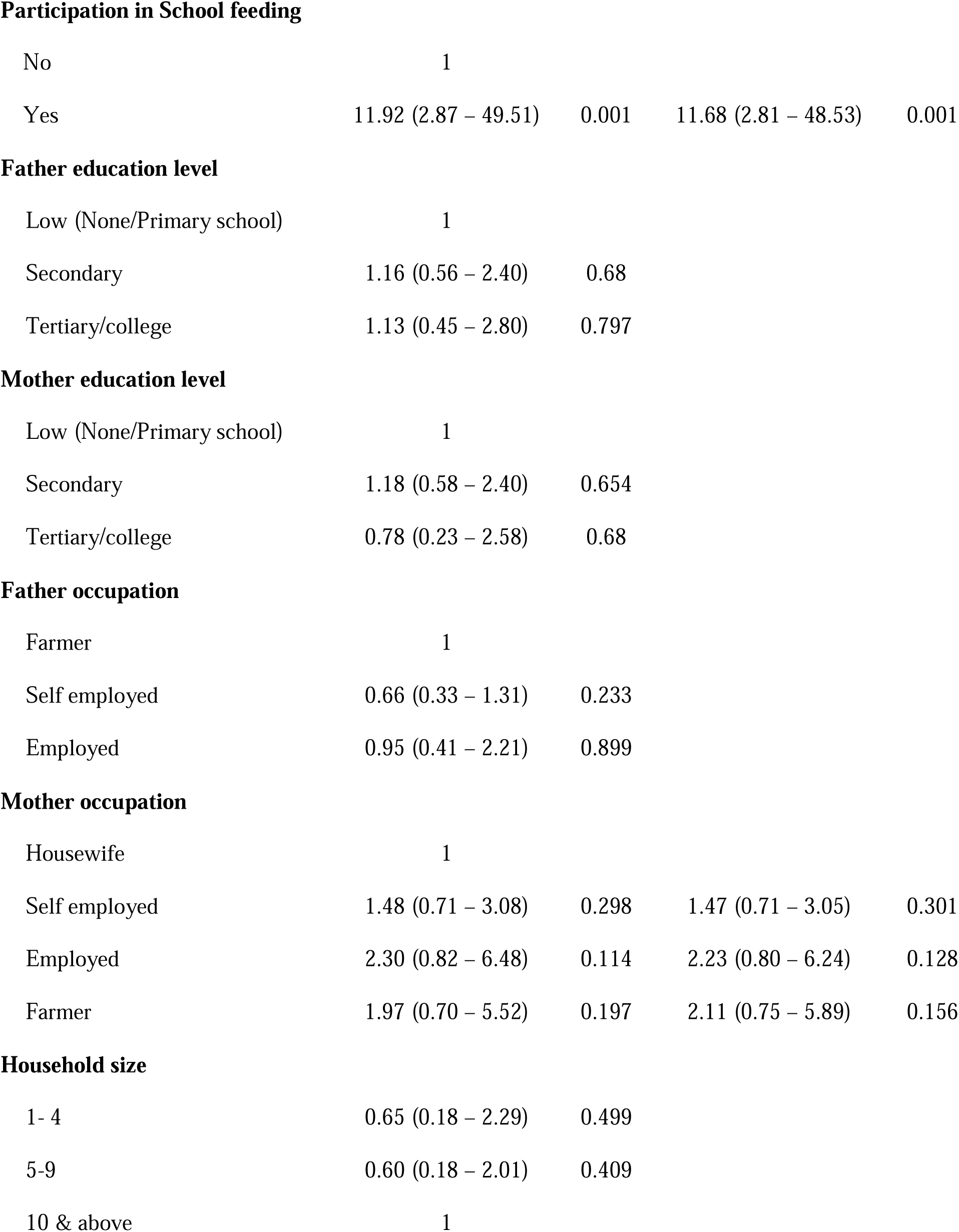
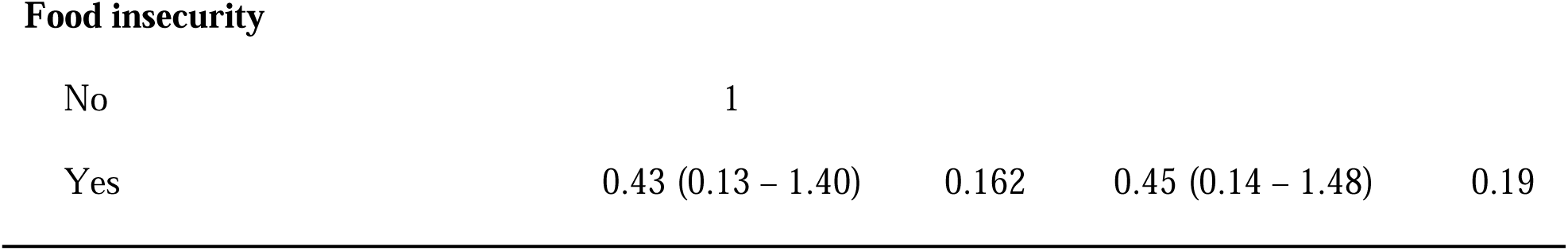
Effect of school feeding program on meal frequency.

| Variable | CPR (95% CI) | *p*-value | APR (95% CI) | *p*-value |
| --- | --- | --- | --- | --- |
| **Gender** |  |  |  |  |
| Female | 1 |  | 1 |  |
| Male | 0.54 (0.29 – 1.02) | 0.057 | 0.52 (0.28 – 0.99) | 0.047 |
| **Participation in School feeding** |  |  |  |  |
| No | 1 |  |  |  |
| Yes | 11.92 (2.87 – 49.51) | 0.001 | 11.68 (2.81 – 48.53) | 0.001 |

**Father education level**
|  |  |  |
| --- | --- | --- |
| Low (None/Primary school) | 1 |  |
| Secondary | 1.16 (0.56 – 2.40) | 0.68 |
| Tertiary/college | 1.13 (0.45 – 2.80) | 0.797 |

**Mother education level**
|  |  |  |
| --- | --- | --- |
| Low (None/Primary school) | 1 |  |
| Secondary | 1.18 (0.58 – 2.40) | 0.654 |
| Tertiary/college | 0.78 (0.23 – 2.58) | 0.68 |

**Father occupation**
|  |  |  |
| --- | --- | --- |
| Farmer | 1 |  |
| Self employed | 0.66 (0.33 – 1.31) | 0.233 |
| Employed | 0.95 (0.41 – 2.21) | 0.899 |
| **Mother occupation** |  |  |  |  |
| Housewife | 1 |  |  |  |
| Self employed | 1.48 (0.71 – 3.08) | 0.298 | 1.47 (0.71 – 3.05) | 0.301 |
| Employed | 2.30 (0.82 – 6.48) | 0.114 | 2.23 (0.80 – 6.24) | 0.128 |
| Farmer | 1.97 (0.70 – 5.52) | 0.197 | 2.11 (0.75 – 5.89) | 0.156 |

**Household size**
|  |  |  |
| --- | --- | --- |
| 1- 4 | 0.65 (0.18 – 2.29) | 0.499 |
| 5-9 | 0.60 (0.18 – 2.01) | 0.409 |
| 10 & above | 1 |  |
| **Food insecurity** |  |  |  |  |
| No | 1 |  |  |  |
| Yes | 0.43 (0.13 – 1.40) | 0.162 | 0.45 (0.14 – 1.48) | 0.19 |

## Discussion

The evaluation of school feeding program implementation in this study revealed considerable gaps in infrastructure and logistical aspects across surveyed schools. Despite the presence of feeding initiatives in all schools assessed, none fully adhered to the standards set out in Tanzania’s National Guidelines for School Feeding Programs. This observation is consistent with findings from a similar study, which noted that only 62.5% of schools in Temeke district had partially implemented the guidelines. Our study further revealed critical infrastructural gaps, most notably the absence of designated dining areas and inadequate facilities for food storage and meal preparation. These deficiencies inevitably compromise the effectiveness and sustainability of the programs. Comparable challenges have been reported in Temeke, where only 37.5% of schools possessed food storage facilities, 25% had functional kitchens, and none provided a cafeteria (21). Taken together, these findings suggest that the barriers to effective school feeding are not isolated to Kisarawe but reflect broader systemic weaknesses in program implementation across different districts.

Moreover, despite the presence of partially implemented feeding programs, only 65% of schoolchildren participated. Nutritional adequacy among participants remained suboptimal, with just 56% achieving minimum dietary diversity and only 15% meeting the recommended feeding frequency. Although participation in school feeding appeared to improve dietary diversity, the association did not reach statistical significance. In contrast, program participation was significantly associated with higher feeding frequency, suggesting some positive but limited impact. Similar studies from Ethiopia have reported stronger effects, with school feeding beneficiaries achieving significantly higher dietary diversity scores, better nutritional status, and improved class attendance compared to non-beneficiaries (17). Likewise, adolescent girls in Sidama region demonstrated improved folate intake, dietary diversity, and meal frequency when enrolled in feeding programs (22). Reviews from sub-Saharan Africa further highlight that home-grown school feeding models can enhance meal diversity and sustainability, especially when linked to local food systems (23). Nonetheless, consistent with global evidence, household food insecurity in our context remained a persistent barrier to achieving both dietary diversity and frequency, underscoring the limits of school-based interventions in isolation.

The implementation of school feeding programs in Kisarawe district was hindered primarily by infrastructural and logistical challenges. Despite these constraints, our findings indicated a positive association between program participation and dietary diversity among students. Comparable barriers have been reported across sub-Saharan Africa, where inadequate infrastructure and limited resources consistently undermine the effective delivery of nutritional interventions and restrict their potential impact. (24,25).

This study provided evidence supporting the positive impact of school feeding programs on meal frequency. Participants enrolled in these programs were significantly more likely to consume the recommended three meals per day compared to non-participants. This aligns with findings from previous studies indicating that school feeding initiatives effectively ensure regular meal consumption, which is critical for maintaining consistent nutrient intake and achieving better nutritional health among children (26,27). The pronounced impact observed in Kisarawe District emphasizes the critical importance of school-based nutrition initiatives in areas where household food insecurity is particularly prevalent.

The observed gender differences in meal frequency, with males being less likely to consume regular meals compared to females, highlight an important consideration for the future development and implementation of school feeding programs. While this study identified this disparity, further exploration is necessary to understand underlying factors, which could include cultural norms, household responsibilities, or social dynamics. Recognizing and addressing such gender disparities through targeted, gender-sensitive strategies is crucial for ensuring equitable access and benefit from nutritional interventions in school settings (28,29).

There is a notable influence of household food insecurity with nutrition adequacy. Households with food insecurity and access can equally affect their children from having recommended diversity and frequency of diets. Although children are protected to the end when hunger strikes, they are equally affected in a context of chronic food insecurity and access. This further emphasizes a strengthened school-based nutrition interventions This finding aligns with previous literature underscoring the need for comprehensive approaches combining school-based interventions with broader socio-economic development initiatives (24,25).

This study had limitations. Reliance on self-reported dietary recall methods introduces potential recall bias, which could affect the accuracy of the reported dietary diversity and meal frequency. To minimize this during the interview, students were asked to mention every food consumed in the previous 24 hours from the time they woke up to the time they went to sleep. The researcher then went further to categorize the food mentioned as required.

### Conclusion

All selected schools in Kisarawe district had school feeding programs in place, yet implementation was hindered by significant infrastructural and logistical challenges. None of the schools fully adhered to the national guidelines, and participation remained limited, with only 65% of pupils enrolled. While participation was positively associated with feeding frequency, the relationship with dietary diversity did not reach a statistical significance level. Household food insecurity further constrained both dietary diversity and feeding frequency, underscoring the deep interconnection between school-based interventions and the broader food security context of families.

Strengthening school feeding programs is therefore not only an educational and health priority but also a national development imperative. These programs serve as a lifeline for children in food-insecure households, supporting nutrition, learning outcomes, and long-term human capital formation. Addressing infrastructural gaps, ensuring reliable food supply chains, and achieving universal inclusion are critical steps to enhance their effectiveness. Aligning these efforts with Tanzania’s commitments to the National Multisectoral Nutrition Action Plan and the Sustainable Development Goals particularly those on zero hunger, health, and education will be essential to transform school feeding into a more equitable and sustainable platform for child well-being and national progress.

## Data Availability

Publicly sharing the raw data would contradict the approved ethical application and the information provided to participants regarding the study and the use of their data. The Institutional Review Board (IRB) requires that a formal data sharing agreement should be in place before releasing data. Therefore, upon request and following the stipulated Tanzanian policies on data sharing, data may be made available to researchers requesting access. Requests can be directed to Muhimbili University of Health and Allied Sciences, Institutional Review Board through

## Acknowledgements

The authors wish to acknowledge dedicated academic staff at the School of Public Health, the Kisarawe District Education Office, head-teachers, students and caregivers for their cooperation, efforts, professional guidance, and unwavering support created a research-friendly environment that was crucial to the conceptualization and implementation of the study.

