## Supplementary material for "Implementation of school-feeding programme and its effect on nutritional adequacy among primary school children in Kisarawe district, Tanzania": SUPPLEMENTARY FILES.pdf

**UNITED REPUBLIC OF TANZANIA**

MINISTRY OF EDUCATION, SCIENCE AND TECHNOLOGY

**OFFICE OF THE DIRECTOR – RESEARCH, PUBLICATIONS AND INNOVATIONS**

Ref. No.DA.282/298/01.C/2693 Date: 11/03/2025

MUHAS-REC-03-2025-2693
JULIETH SEBBA BILAKWATE
SCHOOL OF PUBLIC HEALTH AND SOCIAL SCIENCES
MUHAS

**RE: APPROVAL FOR ETHICAL CLEARANCE FOR A STUDY TITLED: EFFECT OF SCHOOL FEEDING PROGRAM ON NUTRITIONAL ADEQUACY AMONG SCHOOL-AGED CHILDREN IN KISARAWE DISTRICT, TANZANIA.**

Reference is made to the above heading,

I am pleased to inform you that the Chairperson has on behalf of the University Senate, approved ethical clearance of the above-mentioned study, on recommendations of the Senate Research and Publications Committee meeting accordance with MUHAS research policy and Tanzania regulations governing human and animal subjects research.

APPROVAL DATE: 11/03/2025
EXPIRATION DATE OF APPROVAL: 11/03/2026

**STUDY DESCRIPTION:**
**Purpose:**
The purpose of this cross-sectional study is to determine the effect of the implementation of school feeding programs on nutritional adequacy among school-aged children in Kisarawe District, Tanzania.

The approved protocol and procedures for this study is attached and stamped with this letter, and can be found in the link provided: https://irb.muhas.ac.tz/storage/Certificates/Certificate%20-%2016470.pdf and in the MUHAS archives.

**The PI is required to:**

1. Submit bi-annual progress reports and final report upon completion of the study.
2. Report to the IRB any unanticipated problem involving risks to subjects or others including adverse events where applicable.
3. Apply for renewal of approval of ethical clearance one (1) month prior its expiration if the study is not completed at the end of this ethical approval. You may not continue with any research activity beyond the expiration date without the approval of the IRB. Failure to receive approval for continuation before the expiration date will result in automatic termination of the approval for this study on the expiration date.
4. Obtain IRB amendment (s) approval for any changes to any aspect of this study before they can be implemented.
5. Data security is ultimately the responsibility of the investigator.
6. Apply for and obtain Data Transfer Agreement (DTA) from NIMR if data will be transferred to a foreign country.
7. Apply for and obtain Material Transfer Agreement (MTA) from NIMR, if research materials (samples) will be shipped to a foreign country.
8. Apply for and obtain permission to publish the results from National Institute for Medical Research. The application should be before submitting to a publisher and not concurrently through the.
9. Any researcher, who contravenes or fail to comply with these conditions, shall be guilty of an offence and shall be liable on conviction to a fine as per NIMR Act No. 23 of 1979, PART III section 10 (2)
10. The PI is required to ensure that the findings of the study are disseminated to relevant stake holders.
11. PI is required to be versed with necessary laws and regulatory policies that govern research in Tanzania. Some guidance is available on our website https://drp.muhas.ac.tz/.

Dr. Nahya Salim Masoud,
**Chairperson, MUHAS Research and Ethics Committee**

Muhimbili University of Health and Allied Sciences
DIRECTOR
Research, Publications & Innovations
Box 65[illegible]

**ASSENT FORM (English)**

EFFECT OF SCHOOL FEEDING PROGRAMS ON NUTRITIONAL ADEQUACY AMONG SCHOOL AGED CHILDREN IN KISARAWE DISTRICT, PWANI REGION

Participant ID……………….

I am a master's student from Muhimbili University of Health and Allied Sciences conducting research study on effect of school feeding programs of nutritional adequacy among school aged children in Kisarawe District, Pwani Region. I would like to invite you to take part in this study through answering questions in a short interview I will conduct with you. Every detail you will give will be confidential, well preserved and stored. Only the research team will have access to this information. Findings from this study will be disseminated both locally and help improve vaccination services in your community and beyond. Your participation in this study is voluntary and you may choose to stop participating in the study at any time. Your decision to stop participating or refuse to answer particular questions will not affect how you receive vaccination services.

If you have any question about this research or about your role in the study, please feel free to contact me **Julieth Bilakwate** 0714 960988.

Supervisor of the study

Prof. Bruno Sunguya

MUHAS

P.O BOX 65001,

Dar es Salaam

Phone number: +255 713 254 000

Chairperson of Research and Publications Committee

Dr. Nahya Salim

MUHAS.

P.O BOX 65001

Dar es Salaam

I will appreciate if you take part in this research since the information you have is of importance and will help community in general.

Do you agree to participate?

Yes ( ) No ( )

Interviewer Signature……………………

Date of Interview………………………..

**CONSENT (English)**

EFFECT OF SCHOOL FEEDING PROGRAMS ON NUTRITIONAL ADEQUACY AMONG SCHOOL AGED CHILDREN IN KISARAWE DISTRICT, PWANI REGION

Participant ID……………….

I am a master's student from Muhimbili University of Health and Allied Sciences conducting research study on effect of school feeding programs of nutritional adequacy among school aged children in Kisarawe District, Pwani Region. I would like to invite you to take part in this study through answering questions in a short interview I will conduct with you. Every detail you will give will be confidential, well preserved and stored. Only the research team will have access to this information. Findings from this study will be disseminated both locally and help improve vaccination services in your community and beyond. Your participation in this study is voluntary and you may choose to stop participating in the study at any time. Your decision to stop participating or refuse to answer particular questions will not affect how you receive vaccination services.

If you have any question about this research or about your role in the study, please feel free to contact me **Julieth Bilakwate** 0714 960988.

Supervisor of the study

Prof. Bruno Sunguya

MUHAS

P.O BOX 65001,

Dar es Salaam

Phone number: +255 713 254 000

Chairperson of Research and Publications Committee

Dr. Nahya Salim

MUHAS.

P.O BOX 65001

Dar es Salaam

I will appreciate if you take part in this research since the information you have is of importance and will help community in general.

Do you agree to participate?

Yes ( ) No ( )

Interviewer Signature……………………

Date of Interview………………………..

### OBSERVATION CHECKLIST SCHOOL FEEDING PROGRAM

**A1. Name of Observer**

- ◯ 1. Julieth Sebba
- ◯ 2. Frank Kiwango
- ◯ 3. Alfred Secha
- ◯ 4. Samwel Mlawa

**A2. Region**

- ◯ Pwani
- ◯ Option 2

**A3. Ward**

- ◯ 1. Kisarawe
- ◯ 2. Kiluvya
- ◯ 3. Msimbu
- ◯ 4. Masaki

**A4. Name of school**

______________________________________________

**A5. Does the school have a feeding program for students?**

- ◯ 0. No
- ◯ 1. Yes

**B1. Availability of food storage facilities (store)**

- ◯ 0. No
- ◯ 1. Yes

**B2. Availability of cooking facilities (kitchen)**

- ◯ 0. No
- ◯ 1. Yes

**B3. Availability of cafeteria/dining space**

- ◯ 0. No
- ◯ 1. Yes

**B4. Having parents/guardians' food committee**

- 0. No
- 1. Yes

**B5. Proving at least four food groups in a meal**

- 0. No
- 1. Yes

**B6. B6. Provided with food during school hours in the past one academic year.**

- 0. No
- 1. Yes

### DIETARY DIVERSITY AND FOOD FREQUENCY ASSESSMENT FOR SCHOOL-AGED CHILDREN IN KISARAWE DISTRICT, TANZANIA

#### Identification

**A1. Name of interviewer**

- ◯ Julieth Sebba
- ◯ Frank Kiwango
- ◯ Alfred Secha
- ◯ Samwel Mlawa

**A2. Region name**

- ◯ Pwani

**A3. District name**

- ◯ Kisarawe

**A4. Ward name**

- ◯ 1. Kisarawe halmasahuri
- ◯ 2. Kiluvya
- ◯ 3. Msimbu
- ◯ 4. Masaki

**A5. School name**

________________________________________

**A5b. Date of interview**

**A6a. Participant ID**

____________________

#### Demographic Information

**A7. Participant's age (last birthday)**

____________________

**A8. Gender**

- 1. Female
- 2. Male

**A9. Education class**

- 1. Standard 3
- 2. Standard 5
- 3. Standard 6

#### Group

**A10. Father's education level**

- 1. No formal education
- 2. Primary education
- 3. Secondary education
- 4. Tertiary education (College/University)

**A11. Father's occupation**

- 1. Farmer
- 2. Self- employed (eg small business)
- 3. Employed (Part-time/full time)

#### Group

**A12. Mother's education level**

- 1. No formal education
- 2. Primary education
- 3. Secondary education
- 4. Tertiary education (college/university)

**A13. Mother's occupation**

- 1. House wife
- 2. Farmer
- 3. Self employed( eg small business)
- 4. Employed (Part-time/full time)

**A14. Number of people in a household**

______________________

### B.Social economic information

**B1a. What type of fuel does your household use primarily for cooking?**

- ☐ 1. Electricity
- ☐ 2. Gas
- ☐ 3. Kerosene
- ☐ 4. Charcoal
- ☐ 5. Firewood
- ☐ 6. No food is cooked in the household
- ☐ 7. Other, mention

**B1b. Fuel_other**

______________________________

**B2. Do you have the following items at home (select multiple)**

- ☐ 1. Electricity
- ☐ 2. Radio
- ☐ 3. Cellphone
- ☐ 4. Television
- ☐ 5. Refridgerator
- ☐ 6. Computer
- ☐ 7. Camera
- ☐ 8. DVD/CD Player
- ☐ 9. Bed/Mattress
- ☐ 10. Table
- ☐ 11. Chair
- ☐ 12. Cabinet/cupboard
- ☐ 13. Bicycle
- ☐ 14. Motorcycle or scooter
- ☐ 15. Car or truck
- ☐ 16. Solar Panel

### Group

#### » C. Food security

**C1a. In the past 30 days has there ever been no food of any kind in your home due to a lack of resources for food?**

- 0. No
- 1. Yes
- 2. I don't know

**C1b. How many times in the past 30 days has this happened?**

- 1. Rarely (1-2 times)
- 2. Sometimes (3-10 times)
- 3. Often (more than 10 times)
- 4. I don't know

### Group

**C2a. In the past 30 days , did you or anyone in your household fall asleep at night feeling hungry because there was not enough food?**

- 0. No
- 1. Yes
- 3. I don't know

**C2b. How many times in the past 30 days did that happen?**

- 1. Rarely (1-2 times)
- 2. Sometimes (3-10 times)
- 3. Often (more than 10 times)
- 4. I don't know

### Group

**C3a. In the past 30 days did you or anyone in your household go a whole day and night without eating at all because there was not enough food?**

- 0. No
- 1. Yes
- 2. I don't know

**C3b. How many times in the past 30 days did that happen?**

- 1. Rarely (1-2 times)
- 2. Sometimes (3- 10 times)
- 3. Often (more than 10 times)
- 4. I don't know

### Group

#### » D. Dietary Diversity Score

**D1a. Did you eat any of Vitamin A rich fruits and vegetables yesterday eg: mangoes, papaya, pumpkin, carrots, sweet potatoes or other locally available vegetables**

- 0. No
- 1. Yes

**D1b. What time did you eat it?**

- 1. Breakfast
- 2. Breakfast snack
- 3. Lunch
- 4. Lunch snack
- 5. Dinner
- 6. Dinner snack

### Group

**D2a. Did you eat eggs yesterday?**

- 0. No
- 1. Yes

**D2b. What time did you eat it?**

- 1. Breakfast
- 2. Breakfast snack
- 3. Lunch
- 4. Lunch snack
- 5. Dinner
- 6. Dinner snack

### Group

**D3a. Did you eat fish or meat yesterday? eg: beef, pork, lamb, chicken, goat, rabbit, wild game, duck, other birds or fish**

- 0. No
- 1. Yes

#### » Group

**D3b. What time did you eat it?**

- 1. Breakfast
- 2. Breakfast snack
- 3. Lunch
- 4. Lunch snack
- 5. Dinner
- 6. Dinner snack

**D4a. Did you eat, grains, roots and tubers yesterday ? Eg:ugali, potatoes, yams, cassava, or foods made from roots, rice, sorghum**

- 0. No
- 1. Yes

**D4b. What time did you eat it?**

- 1. Breakfast
- 2. Breakfast snack
- 3. Lunch
- 4. Lunch snack
- 5. Dinner
- 6. Dinner snack

### Group

**D5a. Did you eat other vegetables yesterday ? Eg: other vegetables, mchicha, chainise, matembele, sukumawiki and other green leafy vegesgreen leafy vegetables**

- 0. No
- 1. Yes

**D5b. What time did you eat it?**

- 1. Breakfast
- 2. Breakfast snack
- 3. Lunch
- 4. Lunch snack
- 5. Dinner
- 6. Dinner snack

#### Group

**D6a.Did you eat legumes and nuts yesterday? Eg: beans, lentils, grand nuts, cashew nuts, soya and others**

- 0. No
- 1. Yes

**D6b. What time did you eat it?**

- 1. Breakfast
- 2. Breakfast snack
- 3. Lunch
- 4. Lunch snack
- 5. Dinner
- 6. Dinner snack

#### Group

**D7a. Did you eat milk and diary products yesterday? eg: milk, yogurt, cheese**

- 0. No
- 1. Yes

**D7b. What time did you eat it?**

- 1. Breakfast
- 2. Breakfast snack
- 3. Lunch
- 4. Lunch snack
- 5. Dinner
- 6. Dinner snack

**D8a. Did you eat anything (meal or snack) outside of the home yesterday?**

- 0. No
- 1. Yes

#### Group

##### » E. Food frequency Queastionear: Section E.How often you eat at least ONE portion of the following foods and drinks in the previous 24 hours?

**E01. Mixed Porridge (Uji)**

- 0. Never
- 1. Once per day
- 2. 2-3 times per day
- 3. 4 times and above per day

**A6b. RPCM score**

______________________________

**E02. Stiff porridge (Ugali)**

- 0. Never
- 1. Once per day
- 2. 2-3 times per day
- 3. 4 times and above per day

#### Group

**E03. Rice alone**

- 0. Never
- 1. Once per day
- 2. 2-3 times per day
- 3. 4 times and above per day

**E04. Rice and beans**

- 0. Never
- 1. Once per day
- 2. 2-3 times per day
- 3. 4 times and above per day

**E05. Beans alone**

- 0. Never
- 1. Once per day
- 2. 2-3 times per day
- 3. 4 times and above per day

#### Group

**E06. Yogurt**

- 0. Never
- 1. Once per day
- 2. 2-3 times per day
- 3. 4 times and above per day

**E07. Milk**

- 0. Never
- 1. Once per day
- 2. 2-3 times per day
- 3. 4 times and above per day

**E08. Tea with sugar**

- 0. Never
- 1. Once per day
- 2. 2-3 times per day
- 3. 4 times and above per day

#### Group

**E09. Beef**

- 0. Never
- 1. Once per day
- 2. 2-3 times per day
- 3. 4 times and above per day

**E10. Fish**

- 0. Never
- 1. Once per day
- 2. 2-3 times per day
- 3. 4 times and above per day

**E11. Meat soup, mixed dish**

- 0. Never
- 1. Once per day
- 2. 2-3 times per day
- 3. 4 times and above per day

#### Group

**E12. Bread**

- 0. Never
- 1. Once per day
- 2. 2-3 times per day
- 3. 4 times and above per day

**E13. Fruit juice**

- 0. Never
- 1. Once per day
- 2. 2-3 times per day
- 3. 4 times and above per day

**E14. Mango**

- 0. Never
- 1. Once per day
- 2. 2-3 times per day
- 3. 4 times and above per day

#### Group

**E15. Spinach/ Mchicha**

- 0. Never
- 1. Once per day
- 2. 2-3 times per day
- 3. 4 times and above per day

**E16. Fried donut (maandazi)**

- 0. Never
- 1. Once per day
- 2. 2-3 times per day
- 3. 4 times and above per day

**E17. Vitumbua**

- 0. Never
- 1. Once per day
- 2. 2-3 times per day
- 3. 4 times and above per day

#### Group

**E18. Cassava**

- 0. Never
- 1. Once per day
- 2. 2-3 times per day
- 3. 4 times and above per day

**E19. Sweet potato**

- 0. Never
- 1. Once per day
- 2. 2-3 times per day
- 3. 4 times and above per day

**E20. Banana ripe**

- 0. Never
- 1. Once per day
- 2. 2-3 times per day
- 3. 4 times and above per day

#### Group

**E21. Vegetables, mixed dish**

- 0. Never
- 1. Once per day
- 2. 2-3 times per day
- 3. 4 times and above per day

**E22. Cabbage**

- 0. Never
- 1. Once per day
- 2. 2-3 times per day
- 3. 4 times and above per day

**E23. Sardine/dagaa**

- 0. Never
- 1. Once per day
- 2. 2-3 times per day
- 3. 4 times and above per day

#### Group

**E24. Sugary beverages eg: soda**

- 0. Never
- 1. Once per day
- 2. 2-3 times per day
- 3. 4 times and above per day

**E25. Milk tea with sugar**

- 0. Never
- 1. Once per day
- 2. 2-3 times per day
- 3. 4 times and above per day

**E26. Chapati**

- 0. Never
- 1. Once per day
- 2. 2-3 times per day
- 3. 4 times and above per day

**E27. Other wild green vegitables**

- 0. Never
- 1. Once per day
- 2. 2-3 times per day
- 3. 4 times and above per day

**F1. RPCM Score**

______________________________
